# Aligning visual prosthetic development with implantee needs

**DOI:** 10.1101/2024.03.12.24304186

**Authors:** Lucas G. Nadolskis, Lily M. Turkstra, Ebenezer Larnyo, Michael Beyeler

## Abstract

**Purpose:** Visual prosthetics are a promising assistive technology for vision loss, yet research often overlooks the human aspects of this technology. While previous studies focus on the perceptual experiences or attitudes of implant recipients (implantees), *a systematic account of how current implants are being used in everyday life is still lacking*.

**Methods:** We interviewed six recipients of the most widely used visual implants (Argus II and Orion) and six leading researchers in the field. Through thematic analyses, we explored the daily usage of these implants by implantees and compared their responses to the expectations of researchers. We also sought implantees’ input on desired features for future versions, aiming to inform the development of the next generation of implants.

**Results:** Although implants are designed to facilitate various daily activities, we found that implantees use them less frequently than researchers expect. This discrepancy primarily stems from issues with usability and reliability, with implantees finding alternative methods to accomplish tasks, reducing the need to rely on the implant. For future implants, implantees emphasized the desire for improved vision, smart integration, and increased independence.

**Conclusions:** Our study reveals a significant gap between researcher expectations and implantee experiences with visual prostheses. Although limited by access to a small population of implantees, this study highlights the importance of focusing future research on usability and real-world applications.

**Translational relevance:** This retrospective qualitative study advocates for a better alignment between technology development and implantee needs to enhance clinical relevance and practical utility of visual prosthetics.

## Introduction

Visual neuroprostheses, including retinal and cortical implants (commonly known as “bionic eyes”), have shown promise as assistive technology for individuals with blindness^1–7^. These devices, similar to cochlear implants, electrically stimulate remaining neurons in the visual pathway to evoke visual percepts (*phosphenes*)^8,9^. Existing devices have demonstrated improved capabilities in localizing high-contrast objects and aiding basic orientation and mobility tasks^4,10^. Notable examples include Argus II^1^ (Second Sight Medical Products, Inc., Sylmar, CA), the first retinal implant to obtain FDA approval, and its successor, Orion^7^ (Cortigent, Inc., Valencia, CA; formerly Second Sight), a cortical implant that is currently in clinical trials (clinicaltrials.gov: NCT03344848). In addition to neuroprostheses, other promising avenues for sight restoration include optogenetic, gene, and stem cell therapies^11–14^, which offer less invasive alternatives by targeting the genetic and molecular bases of visual impairment.

The Argus II utilizes a 6×10 electrode array implanted on the retina, which receives signals from a camera mounted on glasses to provide visual information (Figure 1A-B). Additionally, the efficacy of the Argus II device is primarily contingent upon the condition of the retina and the electrode-retina distance^15–18^. Some stimulation parameters may enhance the device’s effectiveness, while other factors (e.g., inadvertent activation of passing axon fibers) impose limitations on its performance^17,19,20^. In contrast, the Orion device bypasses the eye altogether, with electrodes implanted directly on the surface of the visual cortex, aiming to restore vision by stimulating the brain’s visual processing areas (Figure 1C). The effectiveness of the Orion varies based on stimulation and neuroanatomical parameters^7,21^ (e.g., amplitude, location, timing). The Argus II has been implanted in 388 recipients worldwide, both commercially and during clinical trials (157 female and 231 male; personal communication with Cortigent, Inc., 2024). The Orion device, still in clinical trials, has been implanted in six recipients (1 female and 5 male), with three remaining implanted to date. There is a notable distinction between clinical trial participants and commercial users regarding selection, training, and ongoing support. Clinical trial volunteers are meticulously selected, receive extensive training, and regularly visit the lab, providing critical feedback for future device iterations. In contrast, commercial users typically receive assistance from third-party companies or centers, often at their own motivation, with less rigorous selection and training protocols, though some may still elect to participate in related research and experiments.

**Figure 1:**
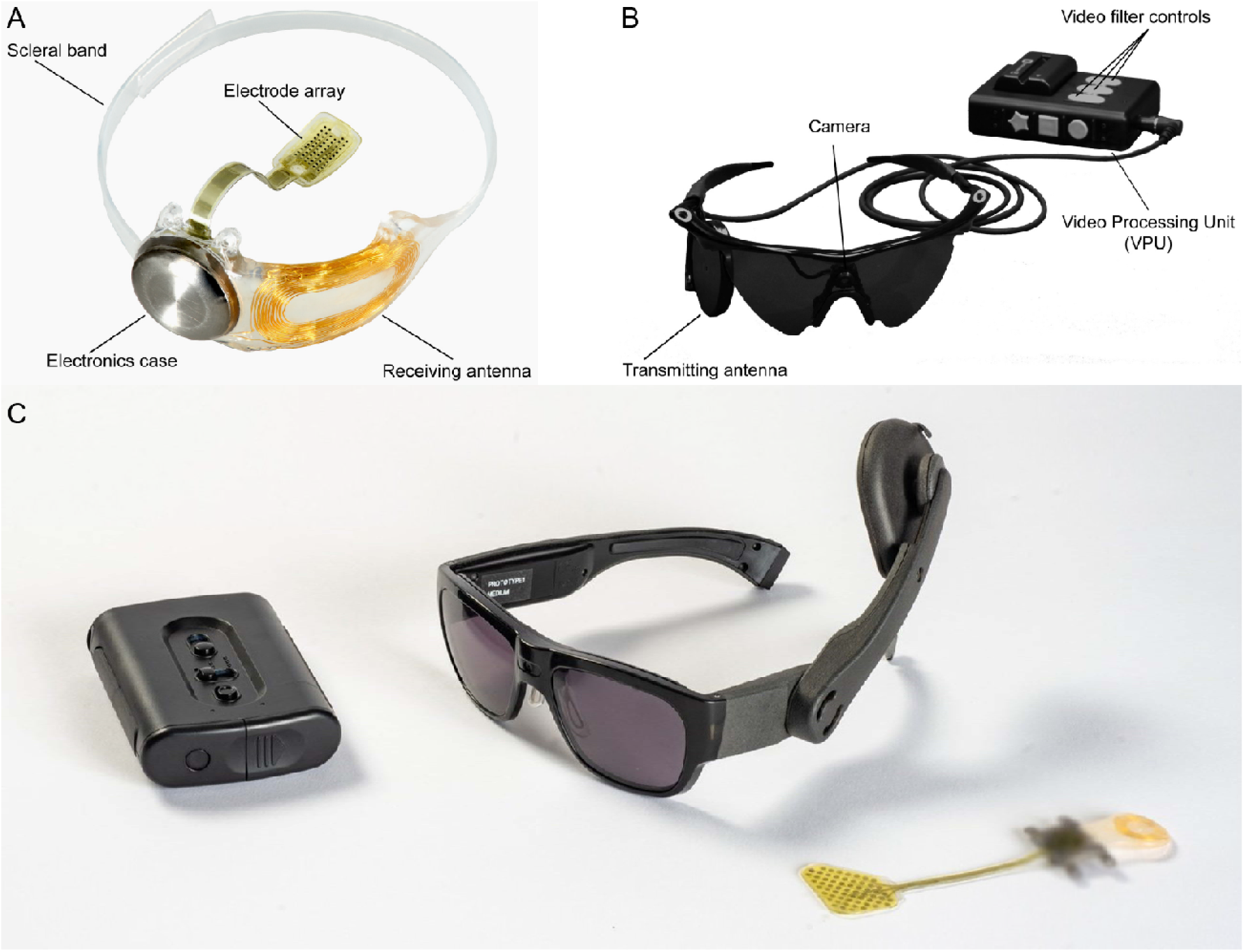
Overview of the Argus II (A-B, reused under CC BY-NC-ND from Ref.^22^) and Orion implants (C, https://cortigent.com). **(A)** The implanted components of the Argus II system include a hermetically sealed enclosure for the electronics that, along with a receiving antenna, is secured to the eye with a scleral band and sutures, and an array of 60 electrodes that is inserted into the eye and tacked over the macula. **(B)** External (body-worn) components of the Argus II system include a pair of glasses with a small camera mounted in the frame connected via a cable to a video processing unit (VPU) worn on the belt or on a shoulder strap. **(C)** This future depiction of the Orion device includes an external data processing unit, which deciphers visual inputs relayed from a miniature camera mounted on a pair of glasses worn by implantees. These inputs are transmitted via electrical pulses to a microelectronic cortical implant situated on the surface of the primary visual cortex. Note: The external components shown represent a future version of the device and were not used by the participants in this study.

Research in artificial vision has traditionally followed the medical model of disability^23^, viewing blindness as a result of an individual’s physical impairment that can be “fixed” – in this case, with an invasive prosthesis. As pointed out by Refs.^24–26^, the majority of research on visual prostheses (and more generally: low vision aids) has primarily focused on technological and functional aspects of these implants (e.g.., the ability to produce phosphenes and the resulting Snellen acuity) and has rarely incorporated implant recipients (*implantees*) in the decision-making and design process^26^. However, blindness is not just about one’s physical impairment, but also about the individual’s subjective psychological experience and the societal contexts in which they live^27,28^. In the development and evaluation of assistive technologies for people who are blind, it is crucial to focus not only on the technical aspects but also on the wants, needs, and lived experiences of the end users, studying how they might utilize such devices within their daily lives. This approach ensures that technology serves the user, enhancing their quality of life rather than solely aiming to correct a physical condition.

Although tools and surveys have been developed to assess the functional visual ability and well-being of implantees^10,29–31^, in practice these are often employed as external validation tools that constitute the very last step of the design process^44,29^. It is therefore perhaps not surprising that none of the current devices have found broad adoption, and that several device manufacturers had to close their doors because their device did not (such as in the case of Retina Implant AG) lead to “the concrete benefit in everyday life of those affected”^1^.

This lack of end-user involvement and limited adoption underscores the necessity for a deeper exploration into how implants are actually used in daily life, contrasted with the initial expectations of their designers. Despite numerous studies assessing functional vision^4,6,29^ and documenting the experiences of current implantees^10,25,32,33^, as well as discussions on ethical considerations in trial participant selection^34,35^ and the attitudes of blind individuals toward implant technology^36^, a comprehensive understanding of the real-world application of these devices remains elusive.

This retrospective qualitative study aims to explore the perspectives, experiences, practices, and aspirations of individuals who have received one of the most commonly available visual implants (Argus II or Orion). It also seeks to contrast these user insights with the viewpoints of prominent researchers who are either involved in developing these devices or who interact directly with the implantees. We also sought feedback from implantees to identify current technology limitations and gather suggestions for future enhancements. Through reconciling the viewpoints of both researchers and implantees as well as fostering cooperative efforts in the design process, we hope that the next generation of visual prosthetic technology can have a profound impact on the quality of life of millions of people worldwide.

## Methods

We conducted semi-structured interviews with 12 participants (six researchers and six implantees, two with the Orion implant and 4 with the Argus II implant) to assess the actual frequency of implant usage among Argus II and Orion users, and compared the reported usage to researcher expectations. This sample represents roughly 1.5% of the global Argus II population and 67% of the individuals who still have the Orion implant, reflecting the rarity of these implants in both commercial and clinical settings. We initially posed structured questions covering an extensive array of instrumental activities of daily living^37,38^ (iADLs). We then engaged in open-ended discussions to explore which strategies and usage patterns that implantees and researchers deemed effective or ineffective, and what the implantees hope to see in the next generation of implants.

The study was deemed exempt from review by the Institutional Review Board of the University of California, Santa Barbara. Participants received an information sheet, ensuring they understood the interview expectations. A minor risk of the study concerned participants feeling embarrassed or experiencing discomfort when answering certain questions during the interview. To mitigate these risks, participants could choose the level of detail they wanted to provide and could choose not to answer specific questions. Researchers also paused the interview if they believed the participant was distressed, confirming whether they wanted to continue. No participants experienced distress or required intervention.

### Study participants

Twelve participants (2 female and 10 male) were recruited via email and phone through a combination of snowball sampling and connections with various research groups and previous collaborators (**Tables 1-2**).

**Table 1.** Participant demographics for each of our interviewed implantees (I1-6). Letters (“A”: Argus II, “O”: Orion) after each participant ID denote the type of implant. “Education” indicates the highest education level completed. Living arrangement: provides a description of other individuals that our participants currently live with, if any.

| Participant | Implant | Age | Gender | Education | Employment status | Years blind | Years implanted | Residential area | Living arrangement | Mobility aid | Top accessibility aids | Cohort |
| --- | --- | --- | --- | --- | --- | --- | --- | --- | --- | --- | --- | --- |
| I1-O | Orion | 25-44 | male | High school | fully employed | 11-20 | 1-5 | city | with family | mobility cane | AI-based smartphone apps, Google maps, voiceover, JAWS | clinical |
| I2-A | Argus II | 45-64 | male | Master's degree | partially employed | 20+ | 6-10 | urban | with family | mobility cane | Voiceover, many different smartphone apps | commercial |
| I3-A | Argus II | 45-64 | male | Bachelor's degree | not employed | 11-20 | 1-5 | urban | with sighted spouse | mobility cane | Screen reader | commercial |
| I4-A | Argus II | 25-44 | male | Associate's degree | fully employed | 20+ | 6-10 | urban | with sighted spouse | guide dog | iPhone with voiceover, navigation apps, social media | commercial |
| I5-O | Orion | 45-64 | male | Bachelor's degree | not employed | 20+ | 6-10 | urban | with family | prefer not to say | Amazon Echo, Siri on phone | clinical |
| I6-A | Argus II | 65+ | female | Bachelor's degree | not employed | 20+ | 6-10 | city | alone | mobility cane, guide dog | iPhone, Android phone, computer with JAWS | commercial |

**Table 2.** Participant demographics for each of our interviewed researchers (R1-6). Letters after each participant ID (“A”: Argus II, “O”: Orion, “AO”: both Argus II and Orion) denote implant(s) worked with. Education level: highest educational qualification, where tertiary may include advanced degrees such as MD or PhD, and secondary may include qualifications like an Associate’s or Bachelor’s degree. Professional background: field in which the researcher is professionally trained. Current focus area: Primary focus of the researcher’s current work, whether it is centered on the implantees or the device. Experience with implantee cohort(s): whether the research has worked with implantees in a clinical-trial or commercial setting. Implantee interaction experience: Types of interactions the researcher has had with implantees. Researchers were not asked about their living situation. None of the researchers were blind.

| Subject | Age | Gender | Education level | Professional background | Years in field | Current focus area | Experience with implantee cohort(s) | Implantee interaction experience |
| --- | --- | --- | --- | --- | --- | --- | --- | --- |
| R1-A | 25-44 | female | tertiary degree | vision science / optometry | 5-10 | implantee-centric | clinical, commercial | patient-centered research |
| R2-AO | 25-44 | male | tertiary degree | engineering | 15-20 | implantee-centric | clinical, commercial | patient-centered research |
| R3-AO | 45-64 | male | tertiary degree | engineering | 15-20 | device-centric, implantee-centric | clinical, commercial | patient-centered research |
| R4-A | 45-64 | male | secondary degree | rehabilitation science | 5-10 | implantee-centric | commercial | clinical interaction |
| R5-O | 25-44 | male | tertiary degree | clinical medicine | 1-5 | device-centric | clinical | clinical interaction |
| R6-AO | 45-64 | male | tertiary degree | engineering | 10-15 | device-centric | clinical, commercial | none |

To qualify for the study, implantees (I1-6) had to be current recipients of either the Argus II or Orion implant. All implantees have had their implant for at least five years, remain currently implanted with their respective devices, and none had reported medical complications with the device. All four Argus II users were part of the commercial cohort (i.e., received their implant after it was FDA-approved in 2013) but have participated in elective research studies. In contrast, the two Orion users are part of the ongoing clinical trial (clinicaltrials.gov: NCT03344848). Five implantees lived with either family or a spouse while one lived alone, and all were frequent users of assistive technology both inside and outside of the home. In addition, four participants reported to be users of either a cane or a guide dog, while one reported using both and one preferred not to answer. All participants resided in either the United States or the Netherlands.

The other six participants (R1-6) were distinguished researchers and medical professionals who are prominent figures in the field of visual neuroprosthetics. These participants included principal investigators and key medical professionals who have played integral roles in the development and clinical application of the Argus II and Orion implants. Each of these researchers has substantial experience and has contributed significantly to the field, ensuring that their insights are both authoritative and relevant. Specifically, R2-AO, R3-AO, and R6-AO had experience with both Argus II and Orion. None of the researchers reported having any visual impairment.

### Interview procedure

The interviews were conducted via video conferencing technology by the two lead researchers of the study, one blind and one sighted. Transcripts were generated using the Otter AI transcription software (Mountain View, CA) and analyzed manually by the research team. Each interview lasted between 30 and 90 minutes.

Three probing interview questions were presented to implantees in order to further understand their experience with their device:

- Q1-I: How often do you use your implant for <iADL>? Choose from: daily, weekly, monthly, yearly, never.
- Q2-I: Please give some examples of how your implant supports <iADL>. What works? What does not?
- Q3-I: What do you wish your implant could do to support/facilitate <iADL>?

These questions were repeated for each of twelve iADLs, drawn from previous literature^37–40^ to ensure a broad spectrum of everyday tasks. These iADLs included essential activities such as meal preparation, housekeeping, transportation, and socializing, chosen for their relevance to independence and quality of life for blind individuals. For each iADL, we formulated a series of questions aimed at uncovering not only the frequency and extent of visual prosthetic use, but also the practical benefits and limitations experienced by implantees. Despite these iADLs often requiring multiple steps for satisfactory completion, we aimed to provide a holistic overview of how individuals in this population conduct these tasks and their overall and individual expectations for their implants in daily activities.

In addition, we sought to understand the discrepancy between the expected and actual use of these devices. Researchers were therefore presented with a similar set of questions, but were asked to reply based on their perception of an implantee’s device usage:

- Q1-R: How often do you expect implantees to use the implant you currently work with for <iADL>? Choose from: daily, weekly, monthly, yearly, never.
- Q2-R: Please give some examples of how the implant you currently work with might support <iADL>.

### Data analysis

To get a qualitative understanding of the device usage for the different iADLs, we performed an inductive thematic analysis^41^ on Q2-I, Q-R, and Q3-I. Themes were individually identified from transcripts, with new ones added for unclassified examples. This iterative process continued across all transcripts until no new themes emerged, culminating in a consensus on 13 definitive themes. Both implantees’ and researchers’ responses were categorized under these themes, with unique codes assigned to each for systematic analysis.

## Results

### Implant usage expectations vs. reported outcomes

**Table 3** provides a detailed breakdown of the responses to Q1-I and Q1-R regarding implant use for various iADLs (see Supplementary Materials for a more quantitative analysis). Overall, the most frequent response from implantees on how often they used their device for specific iADLs was “never,” with I4-A and I6-A reporting no use across any everyday activities. Notably, none of the implantees reported current usage of their implants for meal preparation, reading, managing finances or medication, or using personal electronic devices. However, I3-A stood out as the most frequent user, utilizing his implant daily, particularly for outdoor activities like navigating streets and identifying buildings.

**Table 3.** Implantee (I1-6) reports and researcher (R1-6) perception of implant use frequency for each iADL, using the scale: Daily, Weekly, Monthly, Yearly, or Never.

| Subject | Meal<br>Prep | House-<br>keeping | Laundry | Reading | Shopping | Transp-<br>ortation | Finances | Medi-<br>cation | Personal<br>electronic<br>devices | Social-<br>izing | Hobbies | Employ-<br>ment |
| --- | --- | --- | --- | --- | --- | --- | --- | --- | --- | --- | --- | --- |
| I1-O | Never | Yearly | Monthly | Never | Yearly | Never | Never | Never | Never | Monthly | Yearly | Never |
| I2-A | Never | Never | Never | Never | Never | Monthly | Never | Never | Never | Yearly | Never | Yearly |
| I3-A | Never | Never | Never | Never | Daily | Daily | Never | Never | Never | Daily | Weekly | Daily |
| I4-A | Never | Never | Never | Never | Never | Never | Never | Never | Never | Never | Never | Never |
| I5-O | Never | Never | Never | Never | Never | Yearly | Never | Never | Never | Yearly | Never | Never |
| I6-A | Never | Never | Never | Never | Never | Never | Never | Never | Never | Never | Never | Never |
| R1-A | Daily | Daily | Weekly | Never | Never | Daily | Never | Never | Never | Daily | Monthly | Yearly |
| R2-AO | Never | Never | Weekly | Never | Weekly | Monthly | Never | Never | Never | Daily | Never | Weekly |
| R3-AO | Daily | Daily | Weekly | Monthly | Weekly | Weekly | Yearly | Monthly | Monthly | Daily | Weekly | Daily |
| R4-A | Weekly | Weekly | Weekly | Never | Weekly | Daily | Daily | Never | Never | Daily | Weekly | Daily |
| R5-O | Never | Never | Never | Never | Never | Never | Never | Never | Never | Never | Never | Never |
| R6-AO | Weekly | Weekly | Weekly | Never | Weekly | Daily | Yearly | Daily | Never | Daily | Weekly | Daily |

The researchers, in contrast, had much higher expectations for implant use. Most, except for R5-O, anticipated the implant being used for nearly all iADLs. Researchers in device-centric roles (R3-AO and R6-AO) expected daily or weekly use for most iADLs, particularly in social settings. Despite these expectations, variability in actual use was acknowledged, with socialization emerging as the most likely area for device application according to a majority of researchers.

### Thematic analysis of implant usage

We performed an inductive thematic analysis on Q2-I, Q3-I, Q1-R, and Q-2R (see Methods) to get a qualitative understanding of device usage in daily life, revealing 13 distinct themes that were further categorized into broader topics, as presented in **Table 4**.

**Table 4:**
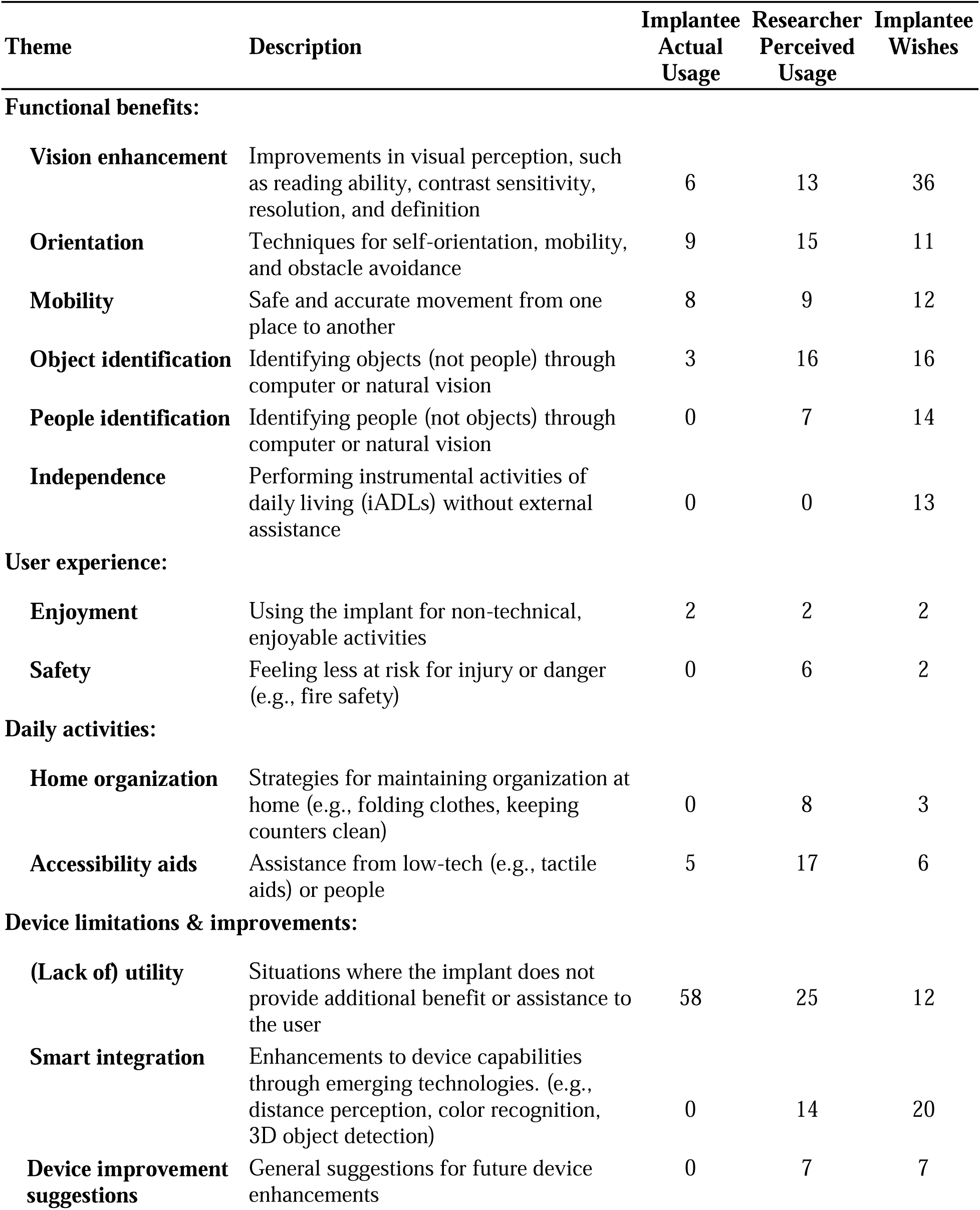
Themes and definitions synthesized from an inductive thematic analysis of interview transcripts. Numbers indicate the counts of themes: of actual implant usage as reported by implantees, of expected implant usage as reported by researchers, of implantee wishes for future visual implants.

Implantees reported *functional benefits* in general visual perception abilities, orientation, and mobility. Researchers perceived these benefits more frequently, with higher numbers in vision enhancement (13), orientation (15), and navigation (9). Implantees expressed a strong wish for future improvements in vision enhancement (36) and navigation (12), indicating a desire for better practical utility in everyday life. Enjoyment and safety were key themes relevant to the *user experience.* While actual usage of the implant for enjoyment was low (2), both researchers and implantees noted the importance of safety, with researchers perceiving more actual usage (6) than implantees reported (0).

In terms of *daily activities*, researchers expected the implant to find widespread use (8), but implantee reports disagreed. Accessibility aids were recognized by both groups, with researchers perceiving more usage (17) compared to implantees’ reports (5).

When discussing *device limitations*, a major theme among implantees was the lack of utility (58), indicating frequent scenarios where the implant did not provide additional benefits. This is somewhat anticipated for tasks like reading and identifying people, where the current implants lack the needed spatial resolution. However, it is less expected for household tasks like housekeeping and meal preparation, areas where one might assume the basic vision enhancement from the implant would offer some advantage. This observation sharply contrasts with researcher expectations (center column), who had anticipated broader application of the implant across a variety of activities.

### Examples of implant usage in the daily life of implantees

To gain deeper insight into everyday device usage, we solicited specific examples (Q2-I and Q2-R) and compared these with researchers’ expectations of device use in daily life. Activities were sorted by their reported frequency of occurrence, as gathered from our interviews, and the most commonly performed activities are discussed below.

Note that some iADLs are more complex than others, often requiring a variety of assistive tools to replace lost visual cues or existing workarounds to complete these tasks without assistive tools. While an implant may not be necessary for all steps of every iADL, our goal is to provide a holistic understanding of how implantees interpret these activities, how they expect implants to aid them, and real-life use cases for their completion.

### Transportation

One application area that both implantees and researchers agree to be of potential value for a visual implant user is that of transportation. Implantees who live in urban areas remarked that the device proved useful for stepping in and out of the bus, avoiding obstacles, and for detecting people when entering and exiting a train. Participant I2-A specifically reported using his implant occasionally to aid him in staying oriented with his surroundings, and to help from bumping into walls and other obstacles. However, the implant could rarely replace the use of a mobility cane or a guide dog completely. In the words of Participant I4-A:

> “I use my device in combination with my guide dog - he walks me to the front of a shop. [Then I use the implant] mostly for orienting myself inside.”

In contrast, Participants I1-O and I6-A, who reside in the city, report that they never use the implant for navigation. Further inquiry revealed that their limited use did not stem from a lack of effort. Specifically, I6-A noted that the ideal scenario for using the implant would be assistance with street-crossing. Yet, the implant’s artificial vision proved too inundating and lacked the necessary detail for her to feel secure using it for navigation in a busy city setting, leading her to prefer being driven as a more reliable alternative. She recounts:

> “I remember my experience crossing the street with the Argus, and that I decided to turn off the stimulation because there were too many flashing lights. The ideal way for me to cross the street is to be able to focus on the important points and detect the distance when crossing…without so much stimulation.”

Participants I1-O, I2-A, and I5-O were quick to remark that the implant slowed them down, as they had learned to (e.g.) navigate much faster and more reliably with a mobility cane. Participant I2-A hoped to use the implant for navigation, but remarked that it did not provide any concrete benefit:

> “I have kind of given up on using it outdoors […], because I hoped it would serve as a navigation device, or something to help me get my bearings and aid in mobility. But it really doesn’t bring any benefit. Lately, I’ve been involved in research studies, just trying to help people understand and advance the science around it, but it doesn’t really provide anything useful enough for me to use in my daily life at all. I don’t know if anybody really does [use it in daily life] at this point.”

### Socializing

Participant I3-A found the implant extremely useful for orientation in social environments, enabling him to monitor the movements of people arriving and departing. This feature was especially helpful for recognizing when someone was approaching to engage in conversation, departing, or coming back, thus helping I3-A discern the presence of others nearby. I3-A mentioned the specific utility of his implant for socialization when navigating through birthday parties or restaurants. I2-A agreed:

> “You know, if you’re sitting at a table, you could maybe tell if somebody was getting up and walking away. Sometimes people have gotten up and walked away while you’re talking, and you end up talking to an empty table. So, there could be some minimal benefit to having the [implant] in a social or entertainment environment.”

Participant I1-O emphasized the implant’s significant role in enhancing social experiences and memorable moments, stating:

> “I use the implant everytime that something new happens to see what I can see. So far I have used it for seeing my birthday candles, fireworks and going to baseball games.”

All researchers similarly emphasized the importance of socialization, and how the implants might facilitate this instrumental aspect of daily life. Various use cases of the implant in socialization settings were mentioned, with R1-A providing some specific context as to how the implant might have aided in a situation similar to that of I2-A above:

> “When people use their implants … they can perceive movements: if someone just left, or if the flash of the person that was right in front of them is not there anymore - they can actually pick that up. [The person in front of them] could be anybody, and they won’t be able to recognize them as their friend or anything, but at least it’s some information.”

### Meal preparation

None of the implantees reported using their devices for meal preparation, although most researchers believed the implant would be somewhat helpful. Implantees mentioned that living with family or a spouse, along with utilizing other assistive tools already in place for cooking, proved more useful for these tasks.

Participant R1-A provided insights into how the device could potentially complement other assistive tools in meal preparation if an implantee chose to use it for this purpose:

> “One aspect of the process for individuals who’ve undergone blind rehabilitation involves having an occupational therapist visit their home to label items and make modifications for easier navigation. This can include adding high-contrast colors to cabinets or using tape and paint for visual cues. With these adaptations in place, introducing an implant can further assist by enhancing their ability to perceive contrasts, helping them locate items by size, and distinguish between things like salt and pepper during meal preparation. Although these improvements might seem basic, they could significantly ease daily activities.”

Other researchers expressed skepticism regarding the device’s suitability for meal preparation, offering more cautious perspectives on its effectiveness as a standalone aid. R6-AO elaborated on these views:

> “The device performs much better when you’re in high-contrast situations, and a kitchen is not necessarily a high-contrast situation. So I would anticipate that this is not the ideal usage situation for a system like this.”

Implantees reported not currently using their devices for meal preparation tasks, primarily due to safety concerns, reliance on existing strategies, and assistance from other tools and cohabitants. I1-O and I4-A highlighted a lack of specific training on utilizing the implant in the kitchen, leading them to prefer established routines. I1-O encapsulated their perspective, stating:

> “I don’t really think I’ve had any success using the device in the kitchen. I can’t even come up with any use cases at the moment.”

### Housekeeping, laundry, and tidying

Regarding housekeeping activities like laundry and maintaining general organization, several implantees noted their attempts to incorporate their implants, albeit with challenges. I5-O had previously tried to use his implant to do laundry, including sorting his socks, but found that:

> “The glasses would actually be more of a problem than a solution - and the cord would get in the way.”

Other implantees found more success when applying their implants to similar tasks. They noted the implant’s utility for specific housekeeping duties, with I1-O highlighting the ability to discern whether lights were on or off, aligning with the device’s effectiveness in high-contrast situations.

Furthermore, researchers had expectations for the implants to facilitate housekeeping activities, especially once an implantee’s home had been customized to suit their needs. R1-A commented:

> “Assuming that their home is already modified to help them with these things… I would think that having an implant would only enhance housekeeping, it can actually enhance the contrast or things of that nature for the objects that they are looking at.”

### Reading

While current implants fall short of enabling the reading of fine print and text, R6-AO challenges the notion of prioritizing reading capabilities in future implant developments, pointing to the superior utility of existing assistive technologies like audiobooks and screen readers. R6-AO stated:

> “I think text-to-voice systems are so advanced that it doesn’t make sense to use the device to read. I see no reason why anybody needs to use the device to read anymore, other than for the pleasure [or joy] of being able to read letters.”

Conversely, R5-O highlighted the desire among potential future implantees to regain some form of reading ability, noting the varied proficiency levels among blind and low-vision individuals with braille and screen readers, and the general wish to read again. Echoing this sentiment, I1-O expressed a specific desire:

> “I would love to be able to read stop signs or road signs. Just having these signs been read to me in some capacity would be great.”

### Other activities of daily living

Despite not being presented as an iADL in our interviews, one common theme between implantees and researchers was the mention of using the device to locate lost, dropped, or missing objects. Participant I1-O specifically mentioned using their implant more frequently to locate their smartphones and computers than to actually operate these personal electronic devices.

However, Participant I3-A (the most prolific implant user in our sample) found less use for the implant for activities that require navigating websites and reading, such as managing his finances. He summarized his thoughts as follows:

> “The reason I no longer use my implant in these different daily activities is because it doesn’t provide a real benefit beyond the techniques that a blind person typically develops to do things.”

Researchers were more positive about the prospects of using the implant they helped design in everyday life. Researchers R1-4, who were more closely involved with the implantees, expected the implant to be used daily or monthly for most iADLs, but also acknowledged that the implant may not be useful at all for some activities. Similarly, R4-A, whose employment focuses on working directly with implantees, states:

> “I think that the blind are already doing many of these kinds of things without assistance. They can do it already in more natural ways, because they have had time to learn and adapt.”

Specifically, these researchers thought that the implant would not be used for reading, managing medication, or using personal electronic devices, but expected the implant to be most commonly used for socializing, transportation, and doing one’s laundry.

Researchers were aware of the current implants’ potential limitations in supporting reading and object recognition. In regard to the employment of implantees, R5-O mentioned that a majority of the employed implantees he works with have daily career tasks tailored to their individual needs, and that an implant might not provide the most aid in these situations.

> “A lot of the subjects that I work with have employment that is adapted to their visual impairment, and in a lot of cases they invested a substantial amount of time and money and effort and retraining to do those.”

When responding to question Q2-R, most researchers referred to recent R&D efforts aimed at enhancing the functionality of the current implant, citing advancements that have not yet been incorporated into the commercial version. Participant R6-AO opted not to comment on the implant’s effectiveness for implantees, citing a disconnect from their experiences:

> “Honestly, this is a question for the users, because as an engineer, as someone who’s working on the device side, I can speak to the performance of the device but I cannot [speak to the user experience side because] I have not been involved on that side of it.”

### Implantee wishes for the next generation of implants

When asked to describe how an ideal future implant would assist in various iADLs (Q3-I and Q3-R), implantees mentioned a wide range of use cases (rightmost column of **Table 4**).

Unsurprisingly, *vision enhancement* topped the list of desires among implantees, aligning with the core promise of bionic eye technologies. This encompasses any improvement in the quality of vision the implant provides. Desired enhancements include better depth perception, as mentioned by I6-A, and color detection. Implantees seemed mostly unaware of ongoing research aiming to improve visual perception through eye movement compensation^42,43^ and stimulus optimization^44–48^, but had heard of efforts to add image filtering^49–51^ and a zoom feature. A prevalent wish among implantees was the ability to read or recognize faces again, though there is a shared understanding that such advancements may not be achievable with current or imminent technology. Reflecting a collective hope, all implantees resonated with I5-O’s desire for any improvement that would enable a shift from relying on tactile to visual cues.

The theme of *smart integration* ranked as the second most frequently mentioned by implantees. All six participants expressed a desire to see their implants work in tandem with widely-used technologies, such as barcode readers, smart glasses, text-to-speech (TTS) audio devices, and color identifiers. Participant I5-O specifically noted the potential benefits of audio enhancements for tasks like meal preparation. Encouragingly, these aspirations align with ongoing research initiatives^49,52–55^. Researchers discussed a variety of smart integration possibilities for different iADLs, including thermal imaging for identifying hot surfaces in the kitchen, as suggested by R3-AO, depth imaging for housekeeping, and compatibility with advanced technological aids like Microsoft’s Seeing AI and OrCam’s MyEye.

*Object identification* emerged as the third most discussed theme, encompassing the ability to discern items ranging from books to debit and credit cards in a wallet, and even bus lines. Researchers believe that current implants already have the potential to facilitate object identification for various iADLs. However, implantees view this capability more as a hope for future device enhancements.

The theme of *independence* stood out among implantees’ aspirations for future implants, yet it was notably absent in discussions about current technology. This discrepancy underscores a gap between the existing capabilities of devices and the ultimate desires of implantees. Participant I5-O expressed a longing for an implant that could assist in securing and maintaining employment by improving his ability to adapt and navigate the workplace.

The prominence of *lack of utility* as a theme highlights the advances that have been made in current assistive technologies and training, indicating that implantees would not need the implant’s assistance to perform certain tasks. However, it also underscores the necessity for future implants to demonstrate their ability to assist people with vision loss in very specific situations. Implantees frequently mentioned this theme when discussing their wishes, expressing that many of their desired functionalities are currently unmet by existing implants. Therefore, a key wish for future implants is to reduce the lack of utility and ensure that the implants provide tangible benefits in a broader range of daily activities.

In essence, implantees envision their implant offering benefits that surpass those provided by traditional mobility aids such as canes, guide dogs, and smartphone applications.

## Discussion

This retrospective qualitative study examines the perspectives of researchers and implantees on the Argus II and Orion visual prostheses. A key finding of our analysis is that socializing emerged as the most frequent use of the implants among implantees, in contrast to other iADLs, which were less frequently supported by the devices. The majority of implantees reported never using their implants for activities such as meal preparation, managing finances, or using personal electronic devices, highlighting a gap between the anticipated use of the implants by researchers and the actual reported use by implantees in everyday life. Researchers, on the other hand, generally expected more frequent use of the implants across most iADLs. This discrepancy emphasizes the importance of aligning device development with real-world user experiences.

### Implant use falls short of researcher expectations

A key finding of our study is that the frequency and application of implant usage did not align with researchers’ expectations. Researchers anticipated that the implants would be used for all iADLs to some extent. However, implantees reported occasional use for specific activities, such as social settings (I1-O, I2-A, I3-A, I5-O), transportation (I2-A, I3-A, I5-O), shopping (I1-O, I3-A), employment (I2-A, I3-A), and around the house (I1-O). Notably, implantees I4-A and I6-A reported never using their implants for everyday activities.

Several factors may account for these discrepancies. First, all four Argus II users were from the commercial cohort and received less support on implant use compared to Orion users, who were part of a clinical trial. This difference in support likely contributed to the more positive experiences reported by Orion users, underscoring the importance of adequate training and support for enabling daily use of these implants. Nevertheless, it is important to note that all four Argus II users had been regularly participating in elective research studies, which provided them with frequent interactions with researchers. This ongoing engagement may have mitigated some of the challenges associated with the reduced initial support. Second, these results should be viewed in light of implantees’ existing skills in navigating blindness before implantation. Participant R2-AO (who had been working with Argus II as well as Orion recipients) commented on the efficiency of pre-developed strategies by implantees with employment, suggesting the implant might not enhance, and could even impede, their performance. This sentiment is encapsulated in R2-AO’s observation:

> “I suspect that [individuals] we enroll in this study, and likely those who opt for the implant later on, will have already undergone extensive training in blindness skills before getting the implant. Consequently, [getting the implant] probably won’t change how they [perform certain iADLs, as it is] easier for them to stick to the […] method they’ve already mastered as blind individuals.”

Third, all but one interviewed implantee lived with a sighted spouse or family member. This may have potentially reduced the utility of the implant in situations where spouses or family members may have aided if needed, or where preexisting assistive heuristics were established.

Yet, not all researchers fully grasp this perspective, as highlighted by R6-AO’s admission of a disconnect from the patient experience due to his engineering role. This gap between device designers and end users underpins a broader issue: the challenge of ensuring that clinical research aligns with the real-world needs of the blind community. This challenge is exacerbated by the fact that all interviewed researchers are sighted, complicating their ability to truly understand the lived experiences of implantees. Participant I4-A felt the strongest about this:

> “Researchers have no clue how it is to be blind, and do not open themselves up to opportunities [to learn about the blind community].”

We also noted a tendency among implantees to blame themselves for the device’s failures (a phenomenon initially reported by Ref.^13^). This is in stark contrast to how researchers and companies often attribute the successes of the device to its technological capabilities. For instance, Participant I2-A felt his challenges were due to both the implant and his own limitations, and I5-O believed his difficulty in using the implant stemmed from his low vision levels, that his “current vision level and abilities are so low that the implant doesn’t work properly.” Successes are celebrated as triumphs of technology, whereas failures are internalized by users as personal deficiencies.

### Implants need to compete with existing technologies

Implants have shown promise in areas like orientation and navigation, where even current aids, such as smartphone apps, fall short. Researchers acknowledge that these implants operate within a technological landscape filled with pre-existing solutions, setting a high bar for new devices to offer distinct advantages.

Therefore, visual implants might be better off focusing on fulfilling specific needs unmet by other technologies. Participant I1-O’s observation, “None of the [accessibility-related smartphone] apps can do what the implant can do, but the implant can’t do what any of the apps can do,” highlights the potential for implants to *complement* rather than compete with existing aids, leveraging their unique capabilities to fill gaps in the current assistive technology ecosystem.

Moreover, visual implants face competition from other vision restoration approaches, such as stem cell therapies and optogenetics. Each of these methods comes with its own set of challenges and potential benefits. For instance, stem cell therapies aim to regenerate damaged retinal cells, while optogenetics involves using light to control neurons that have been genetically modified. Regardless of the method, it is crucial for any field aiming to restore useful vision to consider the wants and needs of their target population.

It is essential to consider user needs comprehensively, irrespective of the treatment type. Our study aims to contribute valuable insights not only to researchers developing bionic eyes but also to app developers, scientists, and ophthalmologists. Understanding user expectations and real-life applications will ensure that these technologies are tailored to enhance the quality of life for individuals with vision loss. By addressing the practical and emotional needs of users, we can foster the development of more effective and user-centered vision restoration solutions.

### Study limitations

We acknowledge that, as with most qualitative retrospective interview studies, hindsight bias is a potential issue. Recent negative headlines about the Argus II may have further influenced perceptions. To mitigate this, we employed several strategies.

First, we interviewed a diverse group of implantees, representing approximately 1.5% of the global Argus II population and 67% of the individuals who still have the Orion implant. However, the small sample size limits the generalizability of our findings, as it reflects only a fraction of the broader population of visual prosthesis users. Despite this, the study offers valuable insights into real-world implant use. We asked participants to provide specific examples to illustrate their opinions, ensuring that their responses were based on concrete experiences rather than retrospective bias. For instance, I2-A stated:

> “I learned in the first few months of using the Argus II that the device was very limited, contrary to what I had seen from ads of people skiing. But I was fortunate to participate in the early trials and test a lot of the new technologies.”

In contrast, I3-A mentioned positive feelings towards the device, saying that the implant met and occasionally exceeded expectations. However, these feelings did not always translate into the measured usefulness of the implant, as reflected across implantee responses in Table 4.

Second, we conducted a thematic analysis where two researchers independently identified common themes across all participants at different stages of their visual prosthetic journey. These insights are crucial for new and existing companies developing future visual prostheses. While hindsight bias is inevitable, the lived experiences of past implantees offer invaluable guidance for the next generation of visual prostheses.

### Future implants: Vision enhancement, smart integration, and independence

Our study highlights implantee wishes for future implant generations, revealing a profound desire for not only enhanced visual perception but also for greater independence. This underscores a crucial need for advancements that go beyond basic navigation aids and aim for a richer, autonomous life experience. Such feedback illuminates the complex dynamics between technological expectations, personal adaptation, and the real challenges of living with vision impairment, pointing out the stark gap in current implant discussions, which rarely touch on the crucial aspect of independence.

The technology is still in its infancy, and implantees recognize this. While current devices meet their expectations, they anticipate substantial progress. Participant I5-O’s expressed:

> “It met my expectations […] We’re at Orion 1 now. Just wait till we get to Orion 15 […] So, the faster and harder you guys work, the quicker we’ll get there.”

However, fulfilling these user wishes requires addressing fundamental technical challenges in the interactions between electrical stimulation and neuronal activity in the visual system. Issues such as electrical current spread and neuronal crosstalk^56–58^, nonselective activation of neurons^59,60^, phosphene persistence and fading^61–63^, lack of eye movement compensation^42,43^ and retinal remodeling^64,65^ significantly influence neural responses to stimulation^19,66–69^. Interviewed implantees were not aware of these technical challenges, but overcoming them will require significant advancements in electrode array design, stimulation protocols, and the integration of emerging technologies. Improved understanding of the neural code of vision may also lead to better strategies for interfacing with the visual system^70^.

Integrating the perspectives and experiences of implantees into the development of future implants may be crucial to transforming this implantable device technology into a vital tool for improving quality of life. By prioritizing implantee experiences and needs, and addressing these fundamental technical challenges, we can ensure that upcoming generations of implants not only push the boundaries of what is technically possible but are genuinely useful in the daily life of people who are blind.

## Data Availability

All data produced in the present study are available upon reasonable request to the authors.

## Footnotes

1 https://web.archive.org/web/20200805082212/ https://www.retina-implant.de/en

## References

1. Luo YH, da Cruz L. The Argus((R)) II Retinal Prosthesis System. Prog Retin Eye Res. 2016 Jan;50:89–107.

2. Palanker D, Le Mer Y, Mohand-Said S, Muqit M, Sahel JA. Photovoltaic Restoration of Central Vision in Atrophic Age-Related Macular Degeneration. Ophthalmology. 2020 Feb 25;

3. Stingl K, Schippert R, Bartz-Schmidt KU, Besch D, Cottriall CL, Edwards TL, et al. Interim Results of a Multicenter Trial with the New Electronic Subretinal Implant Alpha AMS in 15 Patients Blind from Inherited Retinal Degenerations. Front Neurosci [Internet]. 2017 [cited 2020 May 27];11. Available from: https://www.frontiersin.org/articles/10.3389/fnins.2017.00445/full

4. Karapanos L, Abbott CJ, Ayton LN, Kolic M, McGuinness MB, Baglin EK, et al. Functional Vision in the Real-World Environment With a Second-Generation (44-Channel) Suprachoroidal Retinal Prosthesis. Transl Vis Sci Technol. 2021 Aug 12;10(10):7–7.

5. Fernández E, Alfaro A, Soto-Sánchez C, Gonzalez-Lopez P, Lozano AM, Peña S, et al. Visual percepts evoked with an intracortical 96-channel microelectrode array inserted in human occipital cortex. J Clin Invest [Internet]. 2021 Dec 1 [cited 2022 Feb 1];131(23). Available from: https://www.jci.org/articles/view/151331

6. Barry MP, Sadeghi R, Towle VL, Stipp K, Puhov H, Diaz W, et al. Preliminary visual function for the first human with the Intracortical Visual Prosthesis (ICVP). Invest Ophthalmol Vis Sci. 2023 Jun 1;64(8):2842.

7. Beauchamp MS, Oswalt D, Sun P, Foster BL, Magnotti JF, Niketeghad S, et al. Dynamic Stimulation of Visual Cortex Produces Form Vision in Sighted and Blind Humans. Cell. 2020 May 14;181(4):774–783.e5.

8. Fernandez E. Development of visual Neuroprostheses: trends and challenges. Bioelectron Med. 2018 Aug 13;4(1):12.

9. Weiland JD, Walston ST, Humayun MS. Electrical Stimulation of the Retina to Produce Artificial Vision. Annu Rev Vis Sci. 2016;2(1):273–94.

10. Duncan JL, Richards TP, Arditi A, Cruz L da, Dagnelie G, Dorn JD, et al. Improvements in vision-related quality of life in blind patients implanted with the Argus II Epiretinal Prosthesis. Clin Exp Optom. 2017;100(2):144–50.

11. Parnami K, Bhattacharyya A. Current approaches to vision restoration using optogenetic therapy. Front Cell Neurosci. 2023 Aug 16;17:1236826.

12. Hu ML, Edwards TL, O’Hare F, Hickey DG, Wang JH, Liu Z, et al. Gene therapy for inherited retinal diseases: progress and possibilities. Clin Exp Optom. 2021 Mar 2;0(0):1–11.

13. Siy Uy H, Chan PS, Cruz FM. Stem Cell Therapy: a Novel Approach for Vision Restoration in Retinitis Pigmentosa. Med Hypothesis Discov Innov Ophthalmol. 2013;2(2):52–5.

14. McGregor JE. Restoring vision at the fovea. Curr Opin Behav Sci. 2019 Dec 1;30:210–6.

15. Ahuja AK, Yeoh J, Dorn JD, Caspi A, Wuyyuru V, McMahon MJ, et al. Factors Affecting Perceptual Threshold in Argus II Retinal Prosthesis Subjects. Transl Vis Sci Technol. 2013 Apr;2(4):1.

16. Yücel EI, Sadeghi R, Kartha A, Montezuma SR, Dagnelie G, Rokem A, et al. Factors affecting two-point discrimination in Argus II patients. Front Neurosci [Internet]. 2022 [cited 2022 Sep 11];16. Available from: https://www.frontiersin.org/articles/10.3389/fnins.2022.901337

17. Hou Y, Nanduri D, Granley J, Weiland JD, Beyeler M. Axonal stimulation affects the linear summation of single-point perception in three Argus II users. J Neural Eng. 2024 Apr;21(2):026031.

18. Gregori NZ, Callaway NF, Hoeppner C, Yuan A, Rachitskaya A, Feuer W, et al. Retinal Anatomy and Electrode Array Position in Retinitis Pigmentosa Patients After Argus II Implantation: An International Study. Am J Ophthalmol. 2018 Sep 1;193:87–99.

19. Granley J, Beyeler M. A Computational Model of Phosphene Appearance for Epiretinal Prostheses. In: 2021 43rd Annual International Conference of the IEEE Engineering in Medicine Biology Society (EMBC). 2021. p. 4477–81.

20. Beyeler M, Nanduri D, Weiland JD, Rokem A, Boynton GM, Fine I. A model of ganglion axon pathways accounts for percepts elicited by retinal implants. Sci Rep. 2019 Jun 24;9(1):1–16.

21. Barry MP, Armenta Salas M, Patel U, Wuyyuru V, Niketeghad S, Bosking WH, et al. Video-mode percepts are smaller than sums of single-electrode phosphenes with the Orion® visual cortical prosthesis. Invest Ophthalmol Vis Sci. 2020 Jun 10;61(7):927.

22. da Cruz L, Dorn JD, Humayun MS, Dagnelie G, Handa J, Barale PO, et al. Five-Year Safety and Performance Results from the Argus II Retinal Prosthesis System Clinical Trial. Ophthalmology. 2016 Oct 1;123(10):2248–54.

23. Marks D. Models of disability. Disabil Rehabil. 1997 Jan 1;19(3):85–91.

24. Htike HM, Margrain TH, Lai YK, Eslambolchilar P. Ability of Head-Mounted Display Technology to Improve Mobility in People With Low Vision: A Systematic Review. Transl Vis Sci Technol [Internet]. 2020 Sep 24 [cited 2021 Jan 4];9(10). Available from: https://www.ncbi.nlm.nih.gov/pmc/articles/PMC7521174/

25. Erickson-Davis C, Korzybska H. What do blind people “see” with retinal prostheses? Observations and qualitative reports of epiretinal implant users. PLOS ONE. 2021 Feb 10;16(2):e0229189.

26. Kasowski J, Johnson BA, Neydavood R, Akkaraju A, Beyeler M. A systematic review of extended reality (XR) for understanding and augmenting vision loss. J Vis. 2023 May 4;23(5):5.

27. Hogan AJ. Social and medical models of disability and mental health: evolution and renewal. CMAJ Can Med Assoc J. 2019 Jan 7;191(1):E16–8.

28. Gatchel RJ, Peng YB, Peters ML, Fuchs PN, Turk DC. The biopsychosocial approach to chronic pain: scientific advances and future directions. Psychol Bull. 2007 Jul;133(4):581–624.

29. Geruschat DR, Richards TP, Arditi A, da Cruz L, Dagnelie G, Dorn JD, et al. An analysis of observerLrated functional vision in patients implanted with the Argus II Retinal Prosthesis System at three years. Clin Exp Optom. 2016 May;99(3):227–32.

30. Kartha A, Singh RK, Bradley C, Dagnelie G. Self-Reported Visual Ability Versus Task Performance in Individuals With Ultra-Low Vision. Transl Vis Sci Technol. 2023 Oct 17;12(10):14.

31. Ayton LN, Rizzo JF III, Bailey IL, Colenbrander A, Dagnelie G, Geruschat DR, et al. Harmonization of Outcomes and Vision Endpoints in Vision Restoration Trials: Recommendations from the International HOVER Taskforce. Transl Vis Sci Technol. 2020 Jul 16;9(8):25.

32. Lane FJ, Huyck MH, Troyk PR. The Experiences Of Recipients Of A Cortical Visual Prosthesis: A Preliminary Analysis Of Nine Participants Expressed Motivation, Decision-making Process, Risks, And Functional Use Of Phosphenes. Invest Ophthalmol Vis Sci. 2012 Mar 26;53(14):5553.

33. Lane J, Rohan EMF, Sabeti F, Essex RW, Maddess T, Dawel A, et al. Impacts of impaired face perception on social interactions and quality of life in age-related macular degeneration: A qualitative study and new community resources. PloS One. 2018;13(12):e0209218.

34. Xia Y, Peng X, Ren Q. Retinitis pigmentosa patients’ attitudes toward participation in retinal prosthesis trials. Contemp Clin Trials. 2012 Jul 1;33(4):628–32.

35. Xia Y, Ren Q. Ethical Considerations for Volunteer Recruitment of Visual Prosthesis Trials. Sci Eng Ethics. 2013 Sep 1;19(3):1099–106.

36. Karadima V, Pezaris EA, Pezaris JS. Attitudes of potential recipients toward emerging visual prosthesis technologies. Sci Rep. 2023 Jul 6;13(1):10963.

37. Finger RP, Tellis B, Crewe J, Keeffe JE, Ayton LN, Guymer RH. Developing the Impact of Vision Impairment–Very Low Vision (IVI-VLV) Questionnaire as Part of the LoVADA Protocol. Investig Opthalmology Vis Sci. 2014 Oct 1;55(10):6150.

38. Terheyden JH, Fink DJ, Pondorfer SG, Holz FG, Finger RP. Instrumental Activities of Daily Living Tools in Very-Low Vision: Ready for Use in Trials? Pharmaceutics. 2022 Nov;14(11):2435.

39. Owsley C, McGWIN GJ, Sloane ME, Stalvey BT, Wells and J. Timed Instrumental Activities of Daily Living Tasks: Relationship to Visual Function in Older Adults. Optom Vis Sci. 2001 May;78(5):350.

40. Turkstra LM, Van Os A, Bhatia T, Beyeler M. Information Needs and Technology Use for Daily Living Activities at Home by People Who Are Blind [Internet]. 2023 [cited 2023 Jul 22]. Available from: https://arxiv.org/abs/2305.03019v1

41. Abdolrahmani A, Kuber R, Branham SM. “Siri Talks at You”: An Empirical Investigation of Voice-Activated Personal Assistant (VAPA) Usage by Individuals Who Are Blind. In: Proceedings of the 20th International ACM SIGACCESS Conference on Computers and Accessibility [Internet]. New York, NY, USA: Association for Computing Machinery; 2018 [cited 2023 Apr 30]. p. 249–58. (ASSETS ’18). Available from: https://dl.acm.org/doi/10.1145/3234695.3236344

42. Caspi A, Roy A, Wuyyuru V, Rosendall PE, Harper JW, Katyal KD, et al. Eye Movement Control in the Argus II Retinal-Prosthesis Enables Reduced Head Movement and Better Localization Precision. Invest Ophthalmol Vis Sci. 2018 Feb 1;59(2):792–802.

43. Caspi A, Barry MP, Patel UK, Salas MA, Dorn JD, Roy A, et al. Eye movements and the perceived location of phosphenes generated by intracranial primary visual cortex stimulation in the blind. Brain Stimulat. 2021 Jul 1;14(4):851–60.

44. Granley J, Relic L, Beyeler M. Hybrid Neural Autoencoders for Stimulus Encoding in Visual and Other Sensory Neuroprostheses. In: Advances in Neural Information Processing Systems [Internet]. 2022 [cited 2023 Apr 4]. p. 22671–85. Available from: https://papers.nips.cc/paper_files/paper/2022/hash/8e9a6582caa59fda0302349702965171-Abstract-Conference.html

45. Granley J, Fauvel T, Chalk M, Beyeler M. Human-in-the-Loop Optimization for Deep Stimulus Encoding in Visual Prostheses. In: Advances in Neural Information Processing Systems [Internet]. 2023 [cited 2024 Jul 13]. p. 79376–98. Available from: https://proceedings.neurips.cc/paper_files/paper/2023/hash/fb06bc3abcece7b8725a8b83b8fa3632-Abstract-Conference.html

46. Haji Ghaffari D. Improving the Resolution of Prosthetic Vision through Stimulus Parameter Optimization [Internet] [Thesis]. 2021 [cited 2021 Oct 3]. Available from: http://deepblue.lib.umich.edu/handle/2027.42/169970

47. de Ruyter van Steveninck J, Güçlü U, van Wezel R, van Gerven M. End-to-end optimization of prosthetic vision. J Vis. 2022 Feb 28;22(2):20.

48. Leite de Castro DDC, Grayden DB, Meffin H, Spencer M. Neural activity shaping in visual prostheses with deep learning. J Neural Eng. 2024 Jul 10;

49. Sadeghi R, Kartha A, Barry MP, Gibson P, Caspi A, Roy A, et al. Thermal and Distance image filtering improve independent mobility in Argus II retinal implant. J Vis. 2019 Dec 1;19(15):23–23.

50. Sadeghi R, Kartha A, Barry MP, Bradley C, Gibson P, Caspi A, et al. Glow in the dark: Using a heat-sensitive camera for blind individuals with prosthetic vision. Vision Res. 2021 Jul 1;184:23–9.

51. McCarthy C, Walker JG, Lieby P, Scott A, Barnes N. Mobility and low contrast trip hazard avoidance using augmented depth. J Neural Eng. 2014 Nov;12(1):016003.

52. McCarthy C, Walker JG, Lieby P, Scott A, Barnes N. Mobility and low contrast trip hazard avoidance using augmented depth. J Neural Eng. 2014 Nov;12(1):016003.

53. Han N, Srivastava S, Xu A, Klein D, Beyeler M. Deep Learning–Based Scene Simplification for Bionic Vision. In: Augmented Humans Conference 2021 [Internet]. Rovaniemi Finland: ACM; 2021 [cited 2022 Jan 14]. p. 45–54. Available from: https://dl.acm.org/doi/10.1145/3458709.3458982

54. Beyeler M, Sanchez-Garcia M. Towards a Smart Bionic Eye: AI-powered artificial vision for the treatment of incurable blindness. J Neural Eng. 2022 Dec;19(6):063001.

55. Sanchez-Garcia M, Martinez-Cantin R, Guerrero JJ. Semantic and structural image segmentation for prosthetic vision. PLOS ONE. 2020 Jan 29;15(1):e0227677.

56. Wilke RGH, Moghadam GK, Lovell NH, Suaning GJ, Dokos S. Electric crosstalk impairs spatial resolution of multi-electrode arrays in retinal implants. J Neural Eng. 2011 Jun;8(4):046016.

57. Jepson LH, Hottowy P, Mathieson K, Gunning DE, Dąbrowski W, Litke AM, et al. Focal Electrical Stimulation of Major Ganglion Cell Types in the Primate Retina for the Design of Visual Prostheses. J Neurosci. 2013 Apr 24;33(17):7194–205.

58. Madugula SS, Gogliettino AR, Zaidi M, Aggarwal G, Kling A, Shah NP, et al. Focal electrical stimulation of human retinal ganglion cells for vision restoration. J Neural Eng. 2022 Dec 19;19(6).

59. Chang YC, Ghaffari DH, Chow RH, Weiland JD. Stimulation strategies for selective activation of retinal ganglion cell soma and threshold reduction. J Neural Eng. 2019 Feb;16(2):026017.

60. Gaillet V, Cutrone A, Artoni F, Vagni P, Mega Pratiwi A, Romero SA, et al. Spatially selective activation of the visual cortex via intraneural stimulation of the optic nerve. Nat Biomed Eng. 2019 Aug 19;1–14.

61. Pérez Fornos A, Sommerhalder J, da Cruz L, Sahel JA, Mohand-Said S, Hafezi F, et al. Temporal Properties of Visual Perception on Electrical Stimulation of the Retina. Invest Ophthalmol Vis Sci. 2012;53(6):2720–31.

62. Avraham D, Jung JH, Yitzhaky Y, Peli E. Retinal prosthetic vision simulation: temporal aspects. J Neural Eng. 2021 Aug;18(4):0460d9.

63. Hou Y, Pullela L, Su J, Aluru S, Sista S, Lu X, et al. Predicting the Temporal Dynamics of Prosthetic Vision [Internet]. arXiv; 2024 [cited 2024 Jul 17]. Available from: http://arxiv.org/abs/2404.14591

64. Jones BW, Marc RE. Retinal remodeling during retinal degeneration. Exp Eye Res. 2005 Aug;81(2):123–37.

65. Pfeiffer RL, Jones BW. Current perspective on retinal remodeling: Implications for therapeutics. Front Neuroanat [Internet]. 2022 [cited 2023 Feb 8];16. Available from: https://www.frontiersin.org/articles/10.3389/fnana.2022.1099348

66. Beyeler M, Rokem A, Boynton GM, Fine I. Learning to see again: biological constraints on cortical plasticity and the implications for sight restoration technologies. J Neural Eng. 2017 Jun 14;14(5):051003.

67. Avraham D, Yitzhaky Y. Simulating the perceptual effects of electrode–retina distance in prosthetic vision. J Neural Eng [Internet]. 2022 [cited 2022 May 17]; Available from: http://iopscience.iop.org/article/10.1088/1741-2552/ac6f82

68. Guo T, Tsai D, Bai S, Shivdasani M, Muralidharan M, Li L, et al. Insights from Computational Modelling: Selective Stimulation of Retinal Ganglion Cells. In: Makarov SN, Noetscher GM, Nummenmaa A, editors. Brain and Human Body Modeling 2020: Computational Human Models Presented at EMBC 2019 and the BRAIN Initiative® 2019 Meeting [Internet]. Cham: Springer International Publishing; 2021 [cited 2020 Aug 9]. p. 233–47. Available from: 10.1007/978-3-030-45623-8_13

69. Golden JR, Erickson-Davis C, Cottaris NP, Parthasarathy N, Rieke F, Brainard DH, et al. Simulation of visual perception and learning with a retinal prosthesis. J Neural Eng. 2019 Apr;16(2):025003.

70. Abbasi B, Rizzo JF. Advances in Neuroscience, Not Devices, Will Determine the Effectiveness of Visual Prostheses. Semin Ophthalmol. 2021 Mar 18;0(0):1–8.

