## Supplemental Materials for "Aligning visual prosthetic development with implantee needs"

### Supplementary Material

#### Statistical analysis of usage expectations vs. reported outcomes

To allow for a more quantitative comparison between researcher and implantee responses, we converted the responses to Q1-I and Q1-R for each iADL to a numerical five-point scale (0: never, 1: yearly, 2: monthly, 3: weekly, 4: daily; **Figure A1**) and used a linear mixed model to confirm that the answers of implantees and researchers were statistically different. This is a valid transformation, since the original data is ordinal in nature (i.e., “daily” > “weekly”, “weekly” > “monthly”) and our mapping to numerical values preserves this order. The linear mixed model used group (“researcher” vs. “implantee”) as a fixed effect and random intercepts for each subject to account for individual differences. The analysis included 144 observations across 12 unique subjects, with each subject contributing 12 observations. The model was estimated using restricted maximum likelihood (REML) via the statsmodels (v0.14.1) package in Python 3.10. We further performed pairwise Tukey's Honest Significant Difference (HSD) tests for each iADL to test the hypothesis that implantees used their device less frequently than researchers expected.

The group effect (researchers vs. implantees) showed a significant difference in rating values (*z* = 2.756, *p* < 0.01), indicating a strong influence of group membership on the outcome. Specifically, researchers rated the frequency of device usage significantly higher than implantees, with an estimated increase of 1.4 in rating value (e.g., from “yearly” to “weekly”, or from “monthly” to “daily”) as compared to implantees (SE = 0.504, *z* = 2.756, *p* < .01, 95% CI [0.401, 2.377]). The variance attributed to random intercepts for subjects was estimated to be 0.444, suggesting variability in rating values across subjects. The model's convergence was successfully achieved, ensuring the reliability of these estimates.

In pairwise comparisons using Tukey's Honest Significant Difference (HSD) test, researchers were consistently found to have significantly higher frequency ratings compared to implantees (*p* < .05) for meal preparation, housekeeping, laundry, transportation, socializing, and hobbies, with mean differences ranging from 1.8333 to 3.0.

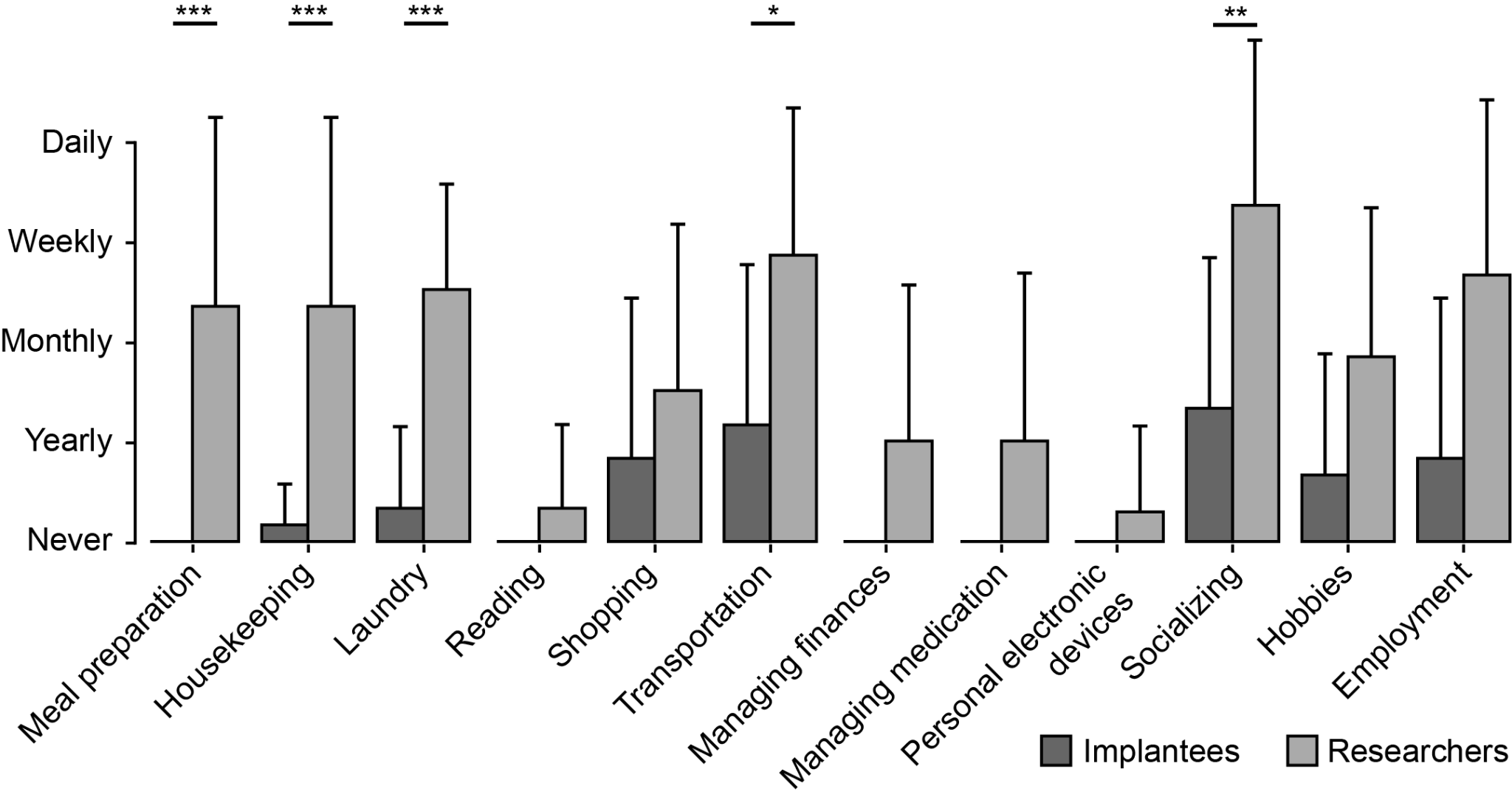

**Figure A1**: Implantee and research perceptions implant use frequency for each iADL. Same data as presented in Table 3, converted to a numerical five-point scale (0: never, 1: yearly, 2: monthly, 3: weekly, 4: daily). Significant differences between implantee and research perceptions (corrected for multiple comparisons using Tukey's Honest Significant Difference test) are denoted as *: p<.05, **: p<.01, ***: p<.001.
